# Genetic analysis of a large heterogeneous patient population with Familial Exudative Vitreoretinopathy

**DOI:** 10.1101/2023.01.11.23284445

**Authors:** Kimberly A. Drenser, Antonio Capone, Matthew Trese

## Abstract

**Purpose:** The purpose of this study is to report the genetic findings of a large heterogeneous patient population (n=551) with the clinical diagnosis of Familial Exudative Vitreoretinopathy (FEVR).

**Methods:** Patients (n=486) were clinically diagnosed with FEVR by exam, birth history, family history, and wide-field fluorescein angiography (WFA). Patients were excluded if WFA was not performed to confirm a proper diagnosis. DNA samples were prospectively collected and analyzed for gene mutations associated with FEVR. The patients represent a heterogenous population: 40% Michigan residents; 48% U.S. residents (non-Michigan); 12% non-U.S. residents. Specifically, alterations in NDP, FZD4, LRP5, TSPAN12, ZNF408, CTNNB1, KIF11 were evaluated.

**Results:** The majority of FEVR patients (69%) had an identifiable gene mutation, with 55% of mutations affecting traditional Wnt-signaling genes: NDP 13%; FZD4 11%; LRP5 25%; TSPAN12 6%. Other affected genes represented 14% of mutations, with ZNF408 accounting for 9%.

**Conclusions:** FEVR remains a clinical diagnosis with WFA necessary for making the diagnosis of FEVR. This database demonstrated that, with proper diagnosis, the majority of patients have a definable genetic alteration.

## Introduction

Familial Exudative Vitreoretinopathy (FEVR) is an inherited condition with multiple genes reported as disease causing. Large databases relying on diagnosis codes have provided interesting findings in regards to incidence and prevalence of affected genes. FEVR, however, remains a clinical diagnosis with a requirement of wide-field fluorescein angiography (WFA).^1,2^ Without WFA, a mis-diagnosis is frequently made, such as a congenital retinal fold or tractional retinal detachment being diagnosed as FEVR. Diagnostic codes are also misleading, as there is no ICD-10 code for FEVR, but rather for “exudative vitreoretinopathy-not diabetes”, which captures a large number of non-FEVR patients.^3^ Associated Retinal Consultants is a tertiary referral center for pediatric retinal vitreoretinopathies and has one of the largest databases of DNA samples for these conditions. The findings of genetic alterations associated with WFA-confirmed FEVR are presented here.

## Methods

Patient DNA samples are prospectively collected through an IRB approved Biobank protocol. Samples linked with the diagnosis of FEVR were evaluated for genetic alterations reported to be associated with FEVR. A diagnosis of FEVR is made by exam, birth history, family history and WFA.^2,3^ If WFA was not obtained to confirm the diagnosis the sample was excluded from analysis. The genes NDP, FZD4, LRP5, TSPAN12, ZNF408, CTNNB1, KIF11 were evaluated. A combination of sanger sequencing, outsourced sequencing and next gen sequencing were used for genetic analysis.

## Results

A previous chart review^3^, revealed that the patients with FEVR examined and/or treated by a pediatric vitreoretinal surgeon at Associated Retinal Consultants are a heterogenous population. Michigan residents account for 40% of patients; U.S. non-Michigan residents account for 48% of patients; non-U.S. citizens account for 12% (19% of non-Michigan residents) of patients.

The Biobank holds 1,356 samples with 551 FEVR samples. Of the 551 samples, 486 underwent genetic testing. The majority (69%) of samples identified a genetic mutation: NDP 13%; FZD4 11%; LRP5 25%; TSPAN12 6%. Genes directly involved in Wnt-signaling account for 55% of mutations identified in FEVR patients. Other affected genes represented 14% of mutations, with ZNF408 accounting for 9%.

## Conclusions

This database represents a heterogenous patient population and suggests that the majority of patients with a clinical diagnosis of FEVR, based on exam, birth history, family history, and WFA have an identifiable mutation. Additionally, the majority of FEVR patients in this database have a mutation affecting Wnt-signaling. Wide-field FA is essential in making the diagnosis of FEVR and greatly increases identifying a genetic association.

## Data Availability

All data produced in the present work are contained in the manuscript

